# Evolutionary dynamics of a virus in a vaccinated population

**DOI:** 10.1101/2021.08.19.21262307

**Authors:** Graham Bell

## Abstract

The progress of an epidemic in a small closed community is simulated by an agent-based model which allows vaccination and variation. The attributes of the virus are governed by two genetic loci: the P-locus, which determines growth, and the M-locus, which determines immune characteristics. Mutation at either locus modifies the attributes of the virus and leads to evolution through natural selection. For both loci the crucial variable is the potential mutation supply U_Pot_, because evolution is likely to happen when U_Pot_ > 1. Mutation at the P-locus causes a limited increase in virulence, which may be affected by vaccine design. Mutation at the M-locus may cause a qualitative shift of dynamic regime from a simple limited epidemic to a perennial endemic disease by giving rise to escape mutants which may themselves mutate. A broad vaccine that remains efficacious despite several mutations at the M-locus prevents this shift and provides protection despite the evolution of the virus. Escape variants may nevertheless arise through recombination after coinfection, and can be suppressed by timely revaccination, using the prevalent strain to design the vaccine.

## Introduction

Once a virus has first infected a single individual in a population of susceptible hosts, its lineage may subsequently follow one of two paths: either it fails to be transmitted and soon becomes extinct, or it proliferates so as to cause an epidemic. Which path is followed depends in part on deterministic factors, such as its initial rate of transmission, and in part on stochastic factors, because the initial survival and transmission of the virus are strongly influenced by chance. The time course of an epidemic in a local population can be adequately described by the mathematical theory that has been developed over the last century (reviewed by Brauer 2017). If the virus is liable to vary when it replicates, however, the virus population will evolve during the epidemic and as its properties change its dynamics may become more complicated (Lenski & May 1994, Day & Proulx 2004, Day & Gandon 2007), as in the well-known case of influenza (reviewed by Nelson & Holmes 2007). As for the virus population as a whole, the spread of any new strain, beginning with a single individual arising by mutation or recombination, will be influenced both by deterministic processes such as natural selection and by stochastic processes such as genetic drift. The ongoing pandemic caused by the coronavirus SARS-CoV-2 has made the public more aware of virus evolution and more fearful of its consequences, as the spread of new strains undermines efforts to control the outbreak (Day et al. 2021).

Novel mutations which arise during the proliferation of an ancestral strain may modify the propensity to infect naïve hosts, which have never before been infected by the ancestor, or the ability to infect recovered hosts, which have previously been infected but then cleared the virus. The first kind, often called virulence or life-history variants, alters the extent of proliferation within the host, which results in a viral titre characteristic of a particular strain. This in turn affects its virulence, because a higher titre causes more severe symptoms and endangers the host, and its transmissibility, because a higher titre results in more virus particles being shed (May & Anderson 1983, van Baalen & Sabelis 1995). The second kind, often called escape variants, alters the antigenic properties of the virus and may thereby enable it to evade the immune system of the host (Gog & Grenfell 2002, Grenfell et al. 2004, Fryer & McLean 2011). These two approaches use different methods and have remained largely separate. The evolution of virulence is usually viewed as a long-term process using quantitative population genetics to model competition between many virus lineages, whereas escape is usually represented as the short-term outcome of competition between two clones of virus (Gandon & Day 2007).

Vaccination protects the population by arming individuals with an immune response against virus strains which have an antigenic profile similar to the vaccine. It is an effective method of reducing the incidence of the disease, and may even drive the virus extinct if the frequency of unvaccinated hosts is too low to sustain an epidemic (Anderson & May 1991). It is also a profound alteration of the host environment, however, and may thereby act as an agent of selection on the virus population. Moreover, vaccinated individuals are not necessarily equivalent to recovered individuals, either in terms of their immune response or the virulence and transmissibility of any infection which succeeds in taking hold. When there are many strains of virus circulating in a vaccinated population, each differing in its virulence, transmissibility and antigenic properties, and each subject to stochastic as well as deterministic change, then analytical solutions of conventional mathematical models based on differential equations may be difficult to find (although Day et al. 2020 show how a complex situation can be analysed). Numerical solutions can still be found, of course, but agent-based simulations can be used instead (e.g. Roche, Drake & Rohani 2011). These can incorporate any number of parameters with ease, at the expense of making it difficult to explore the parameter space thoroughly and correspondingly difficult either to identify or to validate general principles. The most radical approach is to abandon equations altogether, in favour of a set of rules that govern how individuals move around, meet one another, and infect others or are themselves infected. In this report I shall use an agent-based model based on rules rather than equations to describe the evolutionary dynamics of a virus in a vaccinated population. The virus resembles SARS-CoV-2 in some respects, but the model is intended only to suggest some general features of virus evolution, rather than to predict the future course of the pandemic.

## Methods

This report is based on an agent-based, quasi-realistic model of an epidemic in an imaginary small town described in a previous report (Bell 2020). The program itself and a detailed account of the model are included in the Supplementary Material.

### Variation and mutation

The virus has two loci that govern its interaction with a host. The Phenotype or P-locus governs the proliferation of the virus within the host, and thereby its virulence and transmissibility. The Immunity or M-locus governs the interaction of the virus with the immune system of the host. The ancestral strain responsible for the outbreak has given states at the P-locus and the M-locus that determine its transmissibility, virulence and immune characteristics.

Each locus consists of a string of given length of binary digits (‘bits’), one of which is switched from one state to the other when mutation occurs. Competition within the host is neglected, for simplicity, so that only successful mutations are considered – those that both arise and become fixed within an infected host, which then transmits the mutant strain. The effect of a mutation at the P-locus is to increase viral titre by a random factor from the value in the ancestral strain towards some fixed maximal value, leading to an increase, by a different random amount, in both transmissibility and virulence. A mutation at the M-locus alters the antigenic properties of the virus and may thereby enable it to evade the immune system of the host.

The number of mutations at either locus is limited by the number of times that the virus is transmitted, which for the ancestral strain is at most equal to the size of the host population. We can then define the potential mutation supply for either locus as U_Pot_ = HuL, where H is the number of individuals in the host population, u is the fundamental rate of mutation per site, and L is the number of mutable sites at the locus. In an unvaccinated population, at least one mutation will arise, on average, if U_Pot_ > 1. Mutation and the initial spread of a mutant strain are stochastic events, however, so no mutation may occur, or, if it does, the new strain may soon become extinct. In a vaccinated population the condition is more stringent because a successful mutant must arise before the host population is fully vaccinated.

### Immune response

In response to infection by a strain of virus with a given epitope sequence at its M-locus, the host generates an antibody with the complementary sequence and stores this in its immune memory. (For example, if the virus M-locus is 001011001, the host generates and stores 110100110.) If the host survives and is subsequently exposed to the same strain of virus, it is able to express the stored sequence and may thereby disable the virus. The effectiveness of the immune response depends on how the clearance of the virus is related to the degree of complementarity. The simplest mechanism is always to activate the immune response if complementarity equals or exceeds some threshold value, or alternatively the probability of activation might decrease continuously as some function of decreasing complementarity.

### Vaccination

In vaccinated hosts, the sequence complementary to the vaccine sequence is likewise stored in immune memory. Vaccination is akin to previous infection by a virus whose sequence at the M-locus corresponds to the vaccine. It will thereby confer some degree of immunity to this strain of the virus: complete immunity for a highly efficacious vaccine, but only partial immunity for a low-efficacy vaccine. If the host is exposed to a strain of virus whose M-locus does not correspond exactly with the vaccine, its immunity will be less, to an extent that depends on the specificity of the vaccine. A narrow vaccine will activate the immune response, but only if it is fully complementary to an invading virus; a broad vaccine may be less effective against a fully complementary virus, but might be active to some extent against a strain with partial complementarity.

If an individual becomes infected by the virus despite vaccination or previous infection, the virus population within the host will grow, as it does in naïve hosts. It may grow to a different titre in naïve, recovered and vaccinated individuals, however, so that its virulence and transmissibility will in general differ between these categories of host. A weak vaccine permits a higher titre and thereby greater virulence and transmissibility, whereas a strong vaccine suppresses the growth of the virus and reduces both virulence and transmissibility, relative to naïve unvaccinated individuals.

### Standard parameter set

The simulations reported here were conducted in a small town of about 4000 people with a demographic and occupational profile similar to that of many communities in Europe and North America. The parameters for the simulations reported here were chosen so that an epidemic infects about 75 – 80% of an unvaccinated population over a period of about 100 days before the virus is no longer able to propagate and becomes locally extinct. The authorities may take measures to control, curtail or delay the epidemic, including vaccination. A vaccination program proceeds as a specified series of cohorts identified by age and occupation. In the simulations reported here, vaccination gives permanent protection and all individuals are vaccinated during the program, regardless of infection history and current status, although the model allows incomplete compliance and temporary immunity.

## Results and Discussion

### Weak and strong vaccines

The effects of mutation at the P-locus on virulence and transmissibility are antagonistic, because hosts infected by a mutant strain with increased virulence are more likely to die and thus unable to transmit the virus. The spread of a mutant strain is therefore governed by the balance of positive and negative effects on the rate of transmission, which is expected to lead to an optimal intermediate level of virulence (Anderson & May 1982). This balance may be shifted by vaccination. In particular, the cost of virulence arising from the excess death rate of infected hosts is reduced if the virus is able to grow slowly in vaccinated hosts, so that selection will favour increased growth and hence increased virulence. This possibility has led to the controversial conclusion that a weak vaccine may actually favour the spread of more virulent strains of the virus (Gandon et al. 2001; Gandon et al. 2003).

The simplest model is to suppose that naïve hosts are completely susceptible and recovered hosts are completely immune, whereas vaccinated hosts have an intermediate level of immunity. The virulence of the ancestral strain of virus in those vaccinated hosts which become infected is likewise intermediate between its value in naive and recovered hosts. We then expect that greater virulence will evolve in vaccinated hosts, and consequently that virulence in naïve hosts will increase as a correlated response to selection. As mutations at the P-locus arise and spread by virtue of their increased transmissibility, the mean virulence of the virus, over all infected hosts, will tend to increase. The maximum value of mean virulence during the epidemic is shown in Figure 1 for replicate populations with different vaccination states. A weak vaccine does induce the evolution of somewhat greater virulence in naïve hosts, relative to a strong vaccine, but the difference between weak and strong vaccines is not formally significant (F = 3.4; df = 1, 38; 0.1 > P > 0.05), and virulence is greatest in unvaccinated populations because of their greater mutation supply. Selection for increased virulence is expected to be slight, because individuals who die have already had the opportunity to transmit the virus (Day et al. 2020). It is also highly variable because it depends on two stochastic factors: the effect of mutations on virulence and transmissibility, and their timing relative to the vaccination program. The incidence of cases with severe symptoms (which may result in death) is increased by mutation and reduced by vaccination, while the difference between weak and strong vaccines is imperceptible (Supplementary Figure 1).

**Figure 1.**
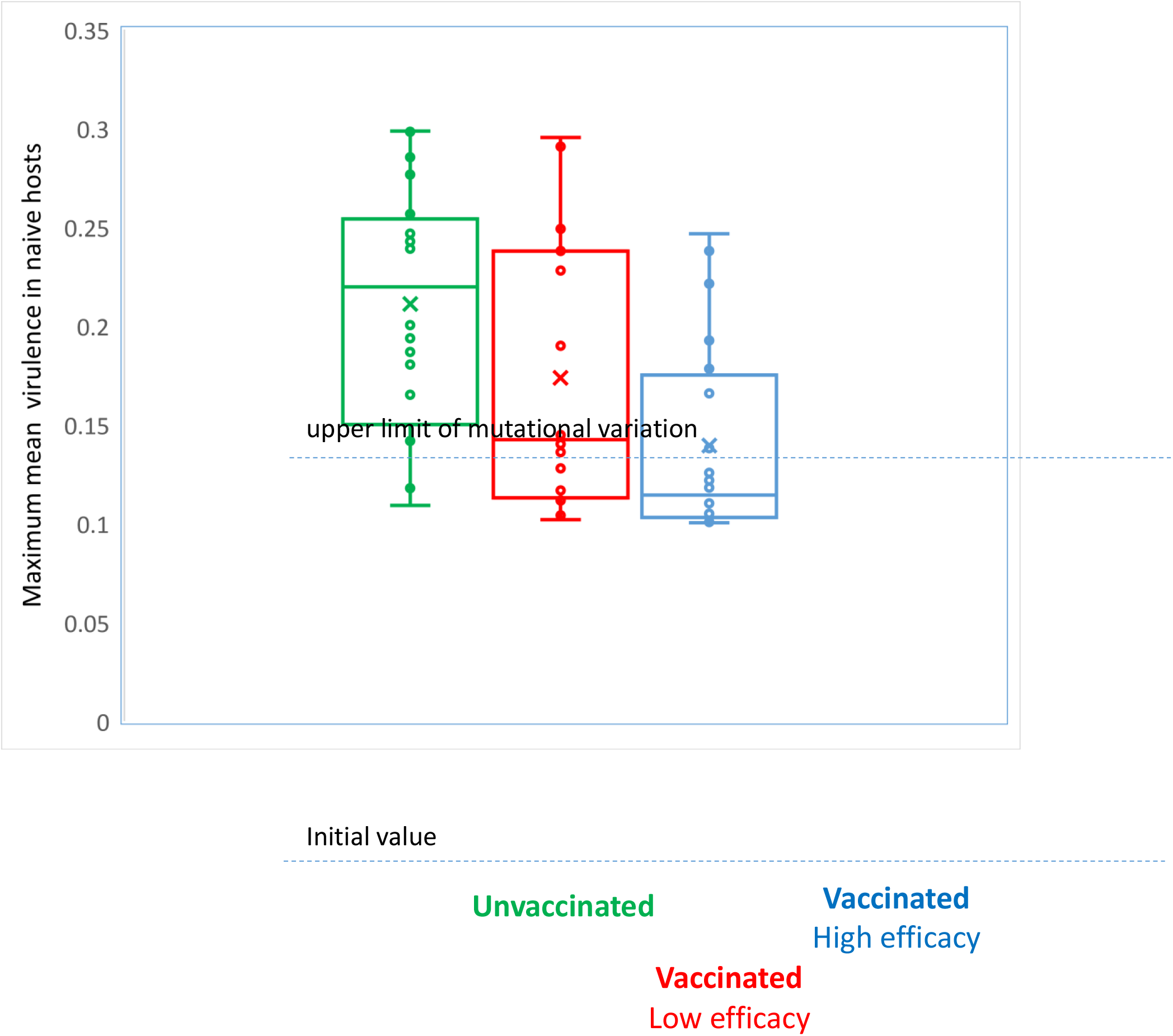
Evolution of virulence in populations with mutation at the P-locus. Plots show the maximum value during the epidemic of mean virulence in naïve (never infected) hosts, for 20 replicate populations with identical initial composition but different vaccination status. The upper limit of variation is set at 3x the initial level of virulence. The values of virulence an

### Evolution of antigenic evasion

Without variation at the M-locus, vaccination using the ancestral strain as a model halts the epidemic and substantially reduces the number of cases, provided that it is efficacious, universally administered and timely. Even if these conditions are met, however, mutations at the M-locus may enable the virus to spread, provided they are sufficiently frequent. If U_Pot_ < 1 it is unlikely (but not impossible) that any mutations will occur, and vaccination will be effective. If U_Pot_ > 1 then escape mutations will usually (although not always) occur, and lineages bearing these mutations can spread even in a fully vaccinated population because they render the whole population susceptible: mutations of this sort create their own potential mutation supply, and may thereby give rise to an indefinite series of further mutations in the future. The succession of mutant strains can shift the population from one dynamic regime to another (Supplementary Figure 2). In the first place, a mutant strain may spread after the initial collapse of the epidemic, which will follow from vaccination or simply from the reduced availability of hosts who are susceptible to the ancestral strain. As the mutant strain in turn declines in abundance it can be replaced by a third mutant strain, if one should occur, and in this way give rise to a more or less regular succession of disease cycles. Alternatively, if U_Pot_ >> 1 mutant strains might arise so frequently that several will be circulating at any given time, and the virus might be maintained for a long period of time with irregular low-amplitude fluctuations over time, before eventually becoming stochastically extinct. Figure 2 shows that the shift from a short-term regular epidemic to a longer-term evolving epidemic in a vaccinated population occurs around the critical potential mutation supply of U_Pot_ ≈ 1. The shift from one dynamic regime to another is thus governed by the potential mutation supply.

**Figure 2.**
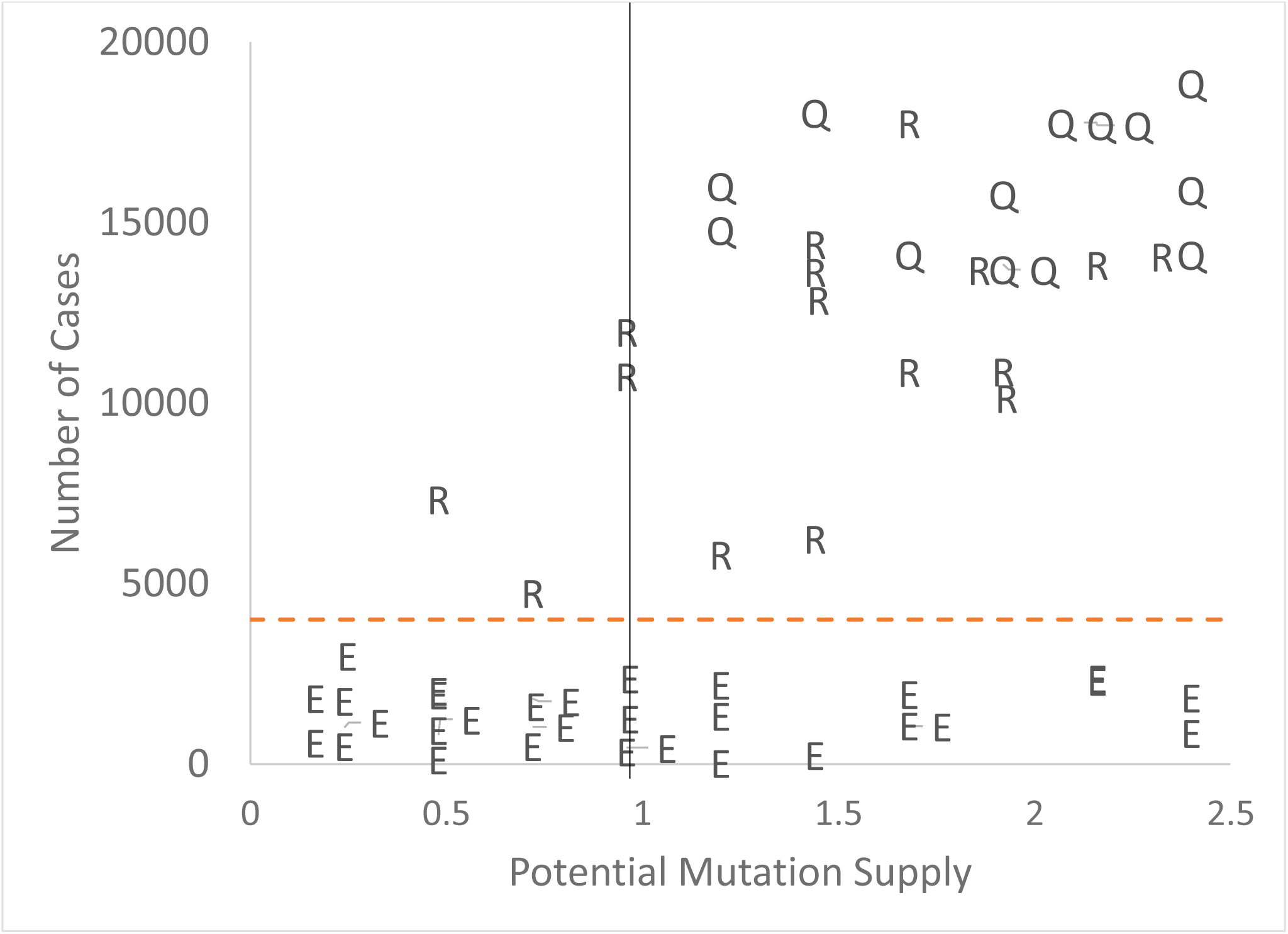
Dynamics of vaccinated populations with different rates of mutation at the M-locus. E denotes a regular single epidemic; R recurrent cyclical epidemics; Q long-term infestation without clear cyclical structure. The x-axis is the potential mutation supply U_Pot_ per locus, with the vertical line showing U_Pot_ ≈ 1 as an average for a vaccinated population in which no mutations occur. The horizontal broken line is the population size. The plot shows 6 replicate populations at each of 10 mutation rates; points have been jiggled slightly for clarity.

No such shift will occur in an unvaccinated population, because escape mutations will spread only in recovered individuals. Most of these will appear relatively early in the epidemic (because the rate of transmission and thus the opportunity for mutation is greatest at this time), before many host individuals have recovered from the infection and are immune to the ancestral strain. Consequently, the density of susceptible hosts is not much greater for a mutant and its selective advantage is only modest, so the lineage will spread only slowly and is likely to die out. In a vaccinated population, on the other hand, the transmission of the ancestral strain is blocked by the vaccine, the selective advantage of mutants is correspondingly much greater, and a mutant lineage can rapidly expand to fill the ecological space provided by vaccinated but susceptible hosts. The potential for vaccination-driven emergence of a vaccine-resistant strain was demonstrated analytically by Scherer & McLean (2002).

The outcome of selection in a vaccinated population will often be a shift in dynamic regime and a substantial increase in the number of cases, relative to a comparable unvaccinated population, because some host individuals will be infected twice, once by the ancestral strain and again by the mutant. If several mutations occur sequentially, each will be likely to spread as its predecessor declines, so that many individuals are infected several times and the total number of cases greatly exceeds that of an unvaccinated population. The effect of vaccination on the total number of cases when there is recurrent mutation at the M-locus is illustrated in Figure 3. In some examples, where no mutation occurs until late in the vaccination program, or fails to spread if it occurs earlier, the resurgence of the epidemic does not occur and vaccination reduces the overall number of infections. In most cases, however, recurrent mutation with U_Pot_ ≈ 1 prolongs the epidemic and causes many more cases than would occur in a comparable unvaccinated population, where the number of cases is limited by the number of host individuals, provided that infection confers immunity in recovered individuals. This surprising result follows from the greater selective advantage of strains which carry a mutation at the M-locus in a vaccinated population.

**Figure 3.**
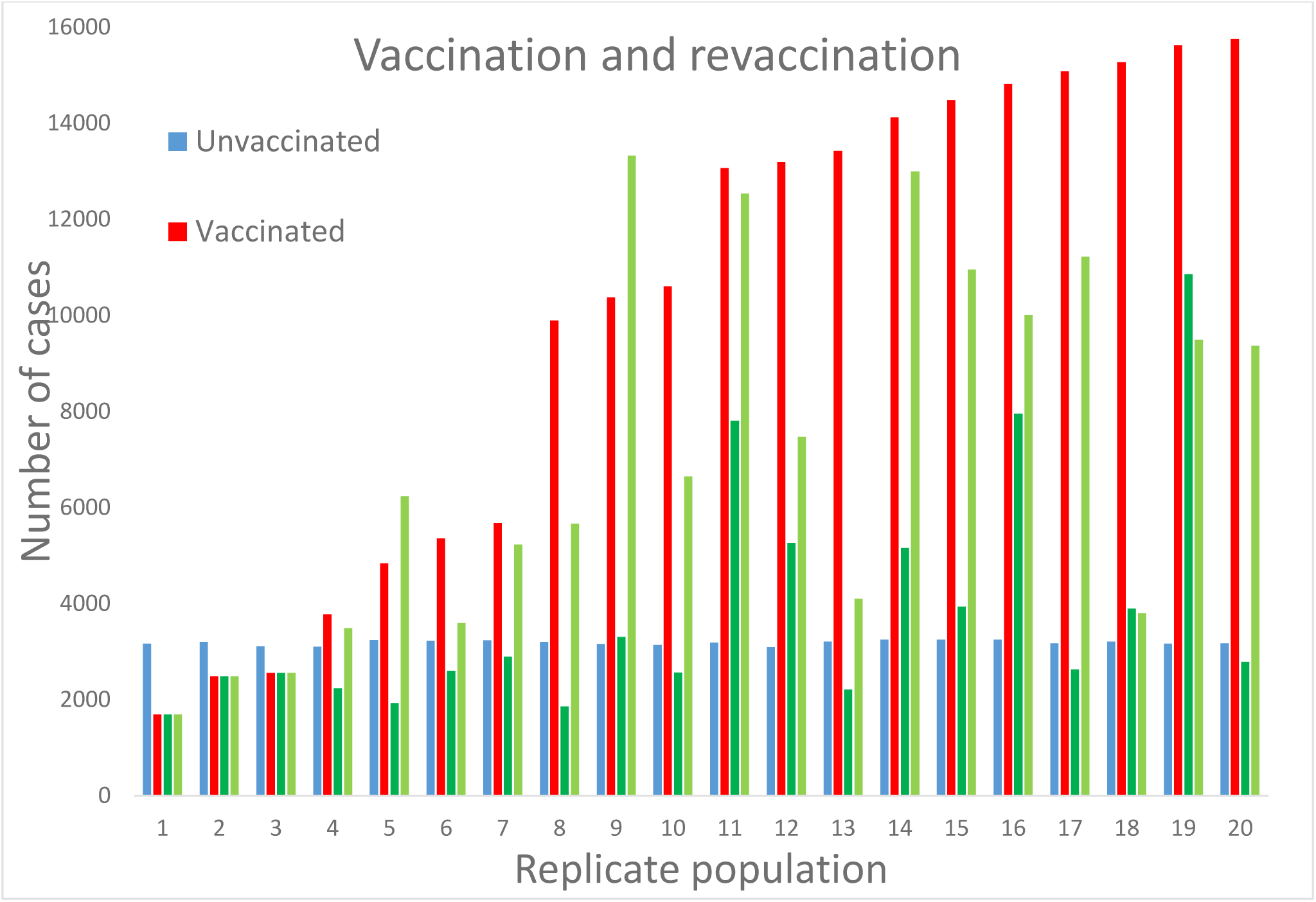
Effect of vaccination and revaccination on the number of cases under recurrent mutation at the M-locus. Treatments for each of 20 replicate populations have identical initial state; different replicates have same parameter values but different initial states. Ranked by number of cases in the vaccinated population. Vaccination beginning 6 weeks after inoculation, in three cohorts spaced by 3 weeks each; revaccination occurs 10 weeks (short lag) or 16 weeks (long lag) after initial vaccination.

### Narrow and broad vaccines

A narrow vaccine is highly specific, being efficacious against the ancestral strain of virus but giving little protection against antigenic variants. A broad vaccine is less active against the ancestral strain, but provides some protection against variants, even when complementarity is incomplete. The perfect vaccine would be efficacious against both the ancestral strain and mutational variants, but efficacy and breadth might be incompatible. Suppose that the efficacy E of a vaccine, the probability that it confers immunity to a given strain of virus, declines exponentially with decreasing complementarity C, such that E_C_ = E_limit_ exp[-k(C_limit_ – C)], where E_limit_ is the efficacy of the vaccine when it corresponds perfectly with the M-locus of the strain (C = C_limit_). A broad vaccine would then have a shallow slope (small k), giving greater protection against similar strains at the expense of giving less protection against matching strains (low E_limit_). Hence, a broad vaccine might reduce the selective advantage of mutations at the M-locus, and thereby prevent the epidemic from becoming cyclical or perpetual; for example, Fryer & McLean (2011) suggested that broad vaccines against HIV might suppress the emergence of escape mutants. This idea was investigated by manipulating E_limit_ and k to produce narrow, medium and broad vaccines in populations where mutation at the M-locus (but not the P-locus) was allowed. The outcome showed little effect of vaccine design, but an overwhelming effect of mutation supply. In unvaccinated populations the epidemic was limited, as before (Figure 4). Vaccinated populations behaved in the same way, provided that few mutations occurred, since most mutations fail to spread. Above a certain threshold, however, escape mutations shifted the population into a new regime with multiple infections per host individual and far more cases overall. A narrow vaccine may predispose the population to shift, but the effect is modest at best (X^2^ = 3.0, df = 1, 0.1 > P > 0.05), whereas the effect of mutation supply is clear.

**Figure 4.**
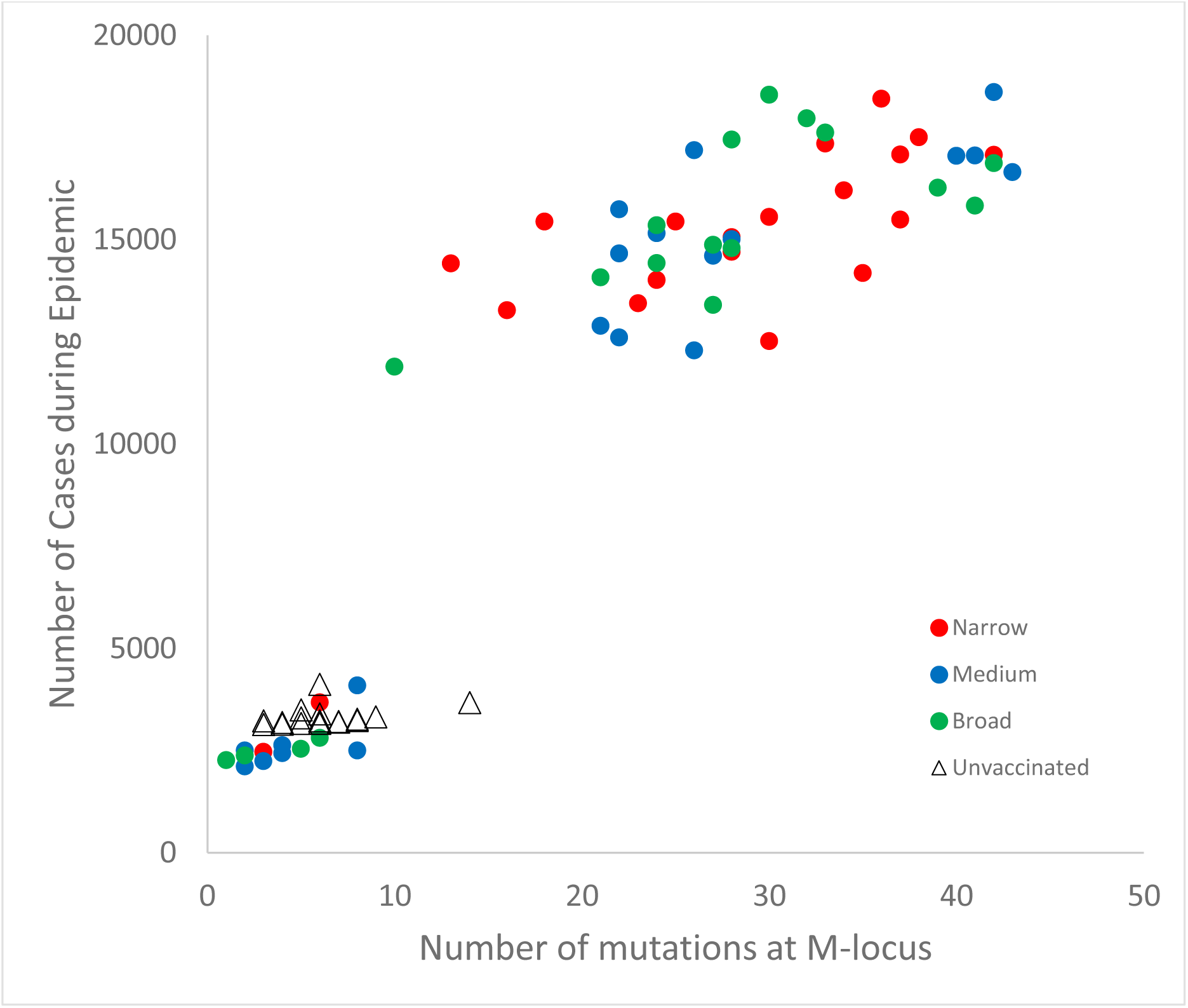
The effect of vaccine design and mutation supply on the number of cases during an epidemic. The plot shows 20 independent replicate populations for each treatment. The x-axis shows all mutations, whether or not they spread.

### Paradox of vaccination

Simple escape mutations at the M-locus not only prolong the epidemic and increase the number of cases, but may shift the dynamic regime from a limited short-term epidemic to a perennial endemic disease. This outcome is not borne out by empirical data on the effects of vaccination. Most vaccines remain effective for long periods of time, and in many cases (e.g. measles) resistance has never evolved. The rarity of vaccine resistance is particularly striking in contrast with the rapid evolution of resistance to antibiotics (Mishra et al. 2012).

This may be a consequence of mutation supply within the host: the immune reaction clears an infection when the pathogen population is still small, whereas antibiotics are usually administered only when the pathogen population has become large enough to cause overt symptoms (Kennedy & Read 2017, 2018; Bell & MacLean 2018). Nevertheless, escape variants have been reported for some vaccines, for example hepatitis B. Even in this case, however, they have failed to eliminate the ancestral strain, perhaps because of a high cost of resistance (Francois et al. 2001). Resistance might often be costly because viral genomes are very compact and hence likely to be disrupted by random change. Reid et al. (2019) reviewed a wide range of models designed to predict the consequences of vaccination against an evolving pathogen, most of which referred to a specific vaccine against a particular pathogen. They concluded that: “Overall, the studies of vaccines that have been in use, have trial data, or have existing homologs predicted positive health outcomes despite vaccine resistance.” The few exceptions require conditions which are thought to be exceptional, such as high levels of cross-immunity between contemporary strains (e.g. Worby et al. 2017). Hence, real-world studies overwhelmingly predict that vaccination will be beneficial, even in the long term against an evolving pathogen.

Vaccine resistance is expected to be rare if the potential mutation supply is usually very low. The rate of mutation per base pair per replication is much higher In RNA viruses than in organisms with a DNA genome, so the requirement that the potential mutation supply be small might seem unlikely. However, the second reason given by Kennedy & Read (2017) for the rarity of vaccine resistance is the extensive redundancy of most vaccines: a variety of antibodies can be produced by the host in response to the presentation of several epitopes on each of several antigens by the pathogen. Consequently, an effective immune response can be mounted against strains similar to, but not identical with, the ancestral strain, leading to the broad cross-reactivity identified by McLean (1995) as a feature of successful vaccines. Most vaccines might therefore tend to be both broad and efficacious, however unlikely such a felicitous combination might seem. Consider a narrow vaccine, which is 100% efficacious against the exactly corresponding sequence at the M-locus of the virus, but otherwise completely inactive, so that a single mutation confers immunity. If the vaccine were somewhat broader, so that it also gave 100% protection against strains that differed at any one of the sites of the M-locus, then a double mutation would be required for immunity. The potential mutation supply now refers to the supply of *double* mutants, which will be proportional to u^2^. The vaccine now restricts the spread of the virus to a single epidemic, and no shift of dynamic regime occurs (Figure 5). This does not require perfect efficacy. If the vaccine is only 70% efficacious, the number of cases is usually increased only slightly, relative to a perfectly efficacious vaccine. On rare occasions, however, a mutation may occur early in the epidemic and drift to high frequency, such that the mutant lineage is liable to undergo a second mutation before the (rather ineffective) vaccination program has eliminated the virus; it is only in this case that a shift in dynamic regime will occur.

**Figure 5.**
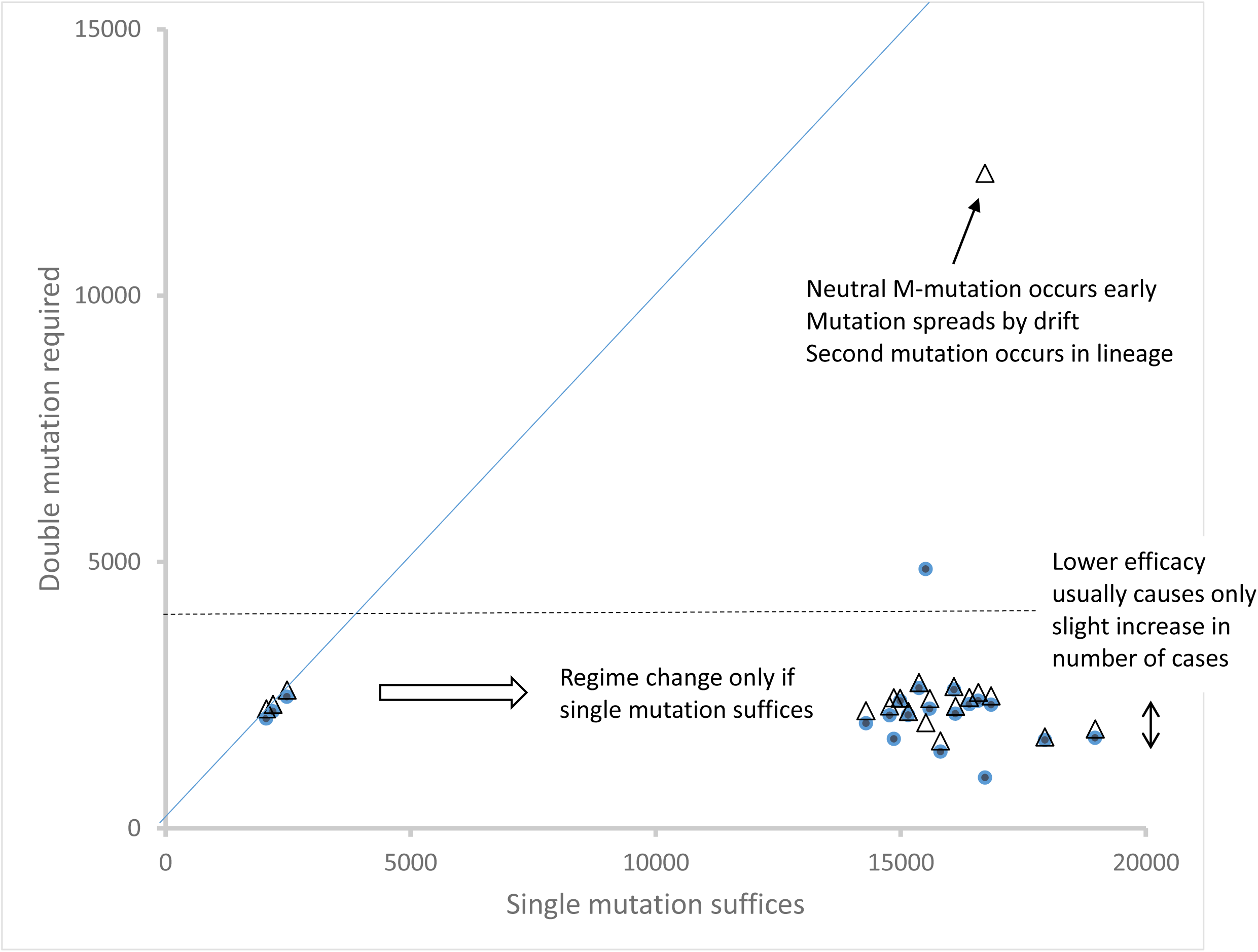
Effect of a broad efficacious vaccine. Individuals are supposed to be immune only if the vaccine is completely complementary to the strain they are exposed to, so that a single mutation at the I-locus of the virus suffices for infection, or that they are immune if the vaccine differs by no more than one site from the strain they are exposed to, so that a double mutation at the M-locus of the virus is required for infection. The values plotted are the total number of cases during the epidemic, for pairs of populations (20 replicate pairs) with identical initial state. Solid circles refer to a vaccine with 100% efficacy, open triangles with 70% efficacy. The solid line is the line of equality. The dashed line marks the approximate population size of 4000 individuals.

### Revaccination

If mutation at the M-locus is generating escape mutants, revaccination might halt the spread of a mutant strain, using the most abundant contemporary strain itself as the model for the vaccine. In an unvaccinated population the epidemic will take its normal course, and die out when the virus is no longer able to proliferate. If a single mutation appears before a vaccination program is fully implemented the ancestral strain is eliminated by the vaccine, producing a fall in the number of new cases, but the mutant strain then spreads rapidly. The outcome is an increase in the total number of cases, as previously noted. Revaccination against the most abundant contemporary strain usually palliates this trend, although it is only effective if administered swiftly (Supplementary Figure 3). Revaccination can reduce the number of excess cases if it is administered soon after the previous vaccination program but if it is delayed its effect is less because the mutant strain has already reached high frequency (Figure 3).

### Recombination

The selection of more highly transmissible variants arising by mutation at the P-locus has only a modest effect on the dynamics of the epidemic, but might be more consequential if they were linked with escape mutants at the M-locus. The probability of stochastic loss of a novel mutation at the M-locus, for example, will be less if it has become linked to a highly transmissible variant caused by mutation at the P-locus. Linkage might arise in two ways: through sequential mutation, or through coinfection leading to intergenic recombination. In principle, recombination is likely to be the more important if coinfection is much more frequent than the rate of mutation per locus. To investigate this possibility, populations with different combinations of mutation and recombination can be set up: neither mutation nor recombination; mutation at the P-locus alone; mutation at the M-locus alone; mutation at both loci without recombination; and mutation at both loci with recombination.

The outcome of the experiment is shown in Figure 6. The first three treatments yield results similar to those already described. With neither mutation nor recombination, the epidemic follows a normal course, infecting a substantial proportion of the population before being knocked down by vaccination and then dying out; mutations at the P-locus which increase virulence and transmissibility cause a modest rise in the number of cases, whereas mutations at the M-locus may lead to a new dynamic regime in which the virus evolves to evade the vaccine, many individuals are infected several times and the total number of cases exceeds the population size. When there is mutation at both loci, but no coinfection, linkage must come about as the consequence of a mutation in one locus occurring in a lineage which already bears a mutation at the other. This causes a modest increase in the average number of cases over the course of the epidemic, relative to a population in which mutation occurs only at the M-locus. If coinfection is permitted, linkage may arise either by sequential mutation or by recombination. This causes a further increase in the average number of cases, suggesting an effect of recombination in addition to the effect of sequential mutation alone. The marginal (additional) effects of mutation and recombination can then be calculated from the five treatment combinations:

**Figure 6.**
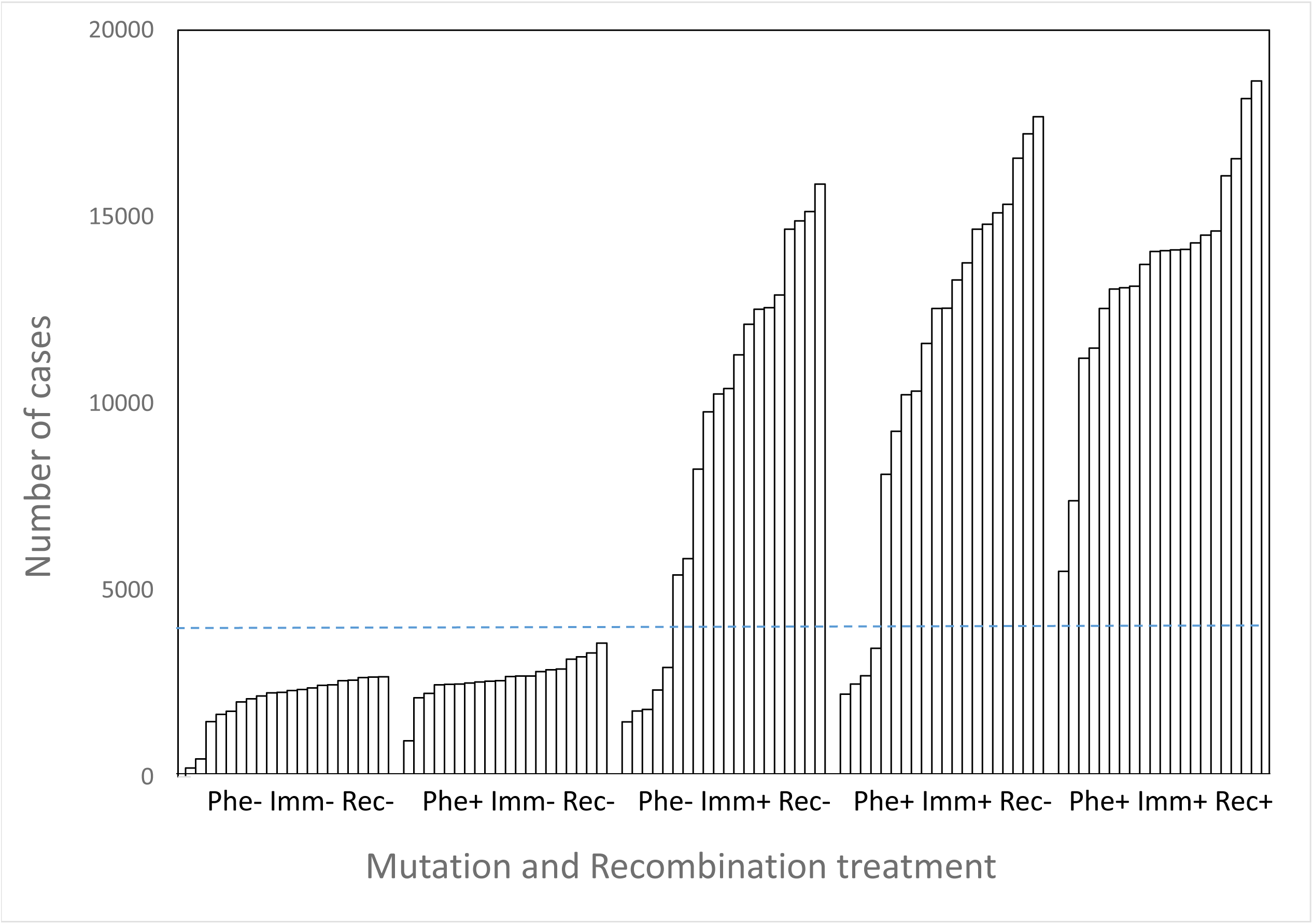
The effect of mutation and recombination on the total number of infections during an epidemic in a vaccinated population. Mutation either occurs or not at the P-locus (with probability 2 × 10^−4^ per bit per replication) and the M-locus (with probability 8 × 10^−5^ per bit per replication), and recombination occurs after coinfection (with probability 0.01 per transmission) or not. Parameters other than the occurrence of mutation or recombination have the same value in all runs. Each of the five treatments has 20 independent replicates, ranked in order of increasing number of cases in each.

|  |  |  |
| --- | --- | --- |
| No mutation or recombination | Phe- Imm- Rec- | scaled to 1 |
| Mutation at P-locus only | Phe+ Imm- Rec- | 1.27 |
| Mutation at M-locus only | Phe- Imm+ Rec- | 4.38 |
| Interaction of P-locus and M-locus | Phe+ Imm+ Rec- | 1.23 |
| Recombination | Phe+ Imm+ Rec+ | 1.21 |

These marginal effects largely depend on whether the dynamic regime is changed by the treatment rather than a smooth increase in the average number of cases. The experiment illustrates the overriding qualitative effect of escape mutations at the M-locus and the modest but appreciable quantitative contribution of mutations at the P-locus, either alone or in combination with mutations at the M-locus. It is possible that the principal effect of recombination, when coinfection is much more frequent than successful mutation, is to facilitate the transition from a regular limited epidemic to a long-term endemic infestation.

## Conclusions

Virus populations will evolve swiftly when there is genetic variation for immune characteristics or within-host growth. The maximum quantity of variation is the potential mutation supply, which is the maximum number of mutations in a simple epidemic: U_Pot_ = HuL. This suggests that there is a threshold at about U_Pot_ = 1, above which mutations are likely to occur, leading to the evolution of the virus population. The simulations based on an agent-based model broadly support the analytical results of previous theory.

Virulence tends to increase when U_Pot_ > 1 for the P-locus, and this may be modulated by vaccine design. The total number of cases during the epidemic cannot exceed the number of host individuals, however, and will usually be substantially less. Any increase in morbidity will be limited, especially when a vaccination program has been implemented.

Mutation at the M-locus may have a qualitatively different outcome. Escape mutations at the M-locus not only prolong the epidemic and increase the number of cases, but may shift the dynamic regime from a limited short-term epidemic to a perennial endemic disease. Both regimes are examples of evolutionary rescue, because a population that would otherwise become extinct is perpetuated by natural selection through the spread of mutants each of which has a positive rate of increase. Simulations confirm that a shift of dynamic regime is likely to occur when the potential mutation supply U_Pot_ > 1. This is because each successful mutation creates its own potential mutation supply and can generate as many new mutations as the ancestral strain. If escape mutations which evade vaccines based on the ancestral virus become established then the epidemic may be prolonged almost indefinitely, unless it can be halted by prompt revaccination. A vaccine which is both broad and efficacious will prevent this shift because only multiply-mutant or recombinant strains will evade the immune response, and it seems likely that most vaccines have this combination of properties.

The classic case of a pathogen that has repeatedly evolved vaccine resistance through mutation and recombination is influenza A virus (reviewed by Taubenberger & Kash 2010). SARS-Cov-2 is also a single-stranded RNA virus, and it is possible that escape mutations will occur. The evolution of SARS-Cov-2 up to February 2021 has been reviewed by Rochman et al. (2021). Altmann, Boyton & Beale (2021) have reviewed the evidence (to March 2021) that mutations affecting the spike protein of SARS-Cov-2 cause changes in viral titre or immune recognition or both. All the variants of concern involve several mutations in the spike protein and must have arisen by sequential mutation or recombination or both. Several variants that have spread recently are resistant to plasma from recovered or vaccinated individuals (Wang et al. 2021), and deletions that cause some degree of resistance arise repeatedly in infected patients (McCarthy et al. 2021). The crucial statistic is the frequency of reinfection, which is poorly defined (but not vanishingly small) for SARS-Cov-2 (Boyton & Altmann 2021).

Although deterministic processes such as mutation supply and natural selection can be recognised, stochastic processes are also important and may be decisive. All the experiments described in this report show a great deal of scatter, with extreme outliers in some cases and qualitatively different outcomes in others, even when the initial state of the population, the mutation rate and the vaccination schedule are identical. The small size of the host population is responsible for some of this scatter, but the initial fate of a mutant is influenced by the small size (initially a single individual) of the mutant lineage itself, independently of the overall size of the population (see Day et al. 2020, Supplementary Material, equation S8), while the schedule and phenotypic effects of mutations are intrinsically stochastic. In some cases, the effect of a treatment (such as vaccination) or a condition (such as mutation) are quite clear, but in others the outcome is dominated by stochastic processes and the consistent effect of an intervention is difficult to discern. Day et al. (2020) make a similar point on the basis of an analytical mathematical model.

More broadly, the dynamics of an epidemic, after the passage of the initial strain, are governed by evolutionary processes involving variation and natural selection. In order to evaluate these processes the most urgent preliminary task is to estimate the potential mutation supply for loci which govern the immune properties and within-host growth of the virus. Practical recommendations which do not include expert guidance on the potential for evolutionary change during the course of an epidemic will be incomplete.

## Supporting information

Detailed information about the model used in the main article.

## Data Availability

This is a theory paper. The program and a detailed description of the model are in the Supplementary Material.

https://www.mcgill.ca/bell-lab/

## Acknowledgments

I am grateful for comments on an earlier version of this article by Austin Burt and Craig MacLean. My research is supported by a Discovery Grant from the Natural Sciences and Engineering Research Board of Canada (Grant RGPIN/6945-2013).

## References

Altmann, D.M., Boyton, R.J. and Beale, R., 2021. Immunity to SARS-CoV-2 variants of concern. Science, 371(6534), pp. 1103–1104.

Anderson, R. M. & May, R. M. 1982 Coevolution of hosts and parasites. Parasitology 85, 411–426.

Anderson, R.M. and May, R.M., 1992. Infectious diseases of humans: dynamics and control. Oxford University Press.

Bell, G. and MacLean, C., 2018. The search for ‘evolution-proof’antibiotics. Trends in Microbiology, 26(6), pp. 471–483.

Benvenuto, D., Giovanetti, M., Ciccozzi, A., Spoto, S., Angeletti, S. and Ciccozzi, M., 2020. The 2019-new coronavirus epidemic: evidence for virus evolution. Journal of medical virology, 92(4), pp. 455–459.

Boyton, R.J. and Altmann, D.M., 2021. Risk of SARS-CoV-2 reinfection after natural infection. The Lancet, 397(10280), pp. 1161–1163.

Brauer, F., 2017. Mathematical epidemiology: Past, present, and future. Infectious Disease Modelling, 2(2), pp. 113–127.

Day, T. and Gandon, S., 2007. Applying population-genetic models in theoretical evolutionary epidemiology. Ecology Letters, 10(10), pp. 876–888.

Day, T., Gandon, S., Lion, S. and Otto, S.P., 2020. On the evolutionary epidemiology of SARS-CoV-2. Current Biology, 30(15), pp. R849–R857.

Day, T. and Proulx, S.R., 2004. A general theory for the evolutionary dynamics of virulence. The American Naturalist, 163(4), pp. E40–E63.

Francois, G., Kew, M., Van Damme, P., Mphahlele, M.J. and Meheus, A., 2001. Mutant hepatitis B viruses: a matter of academic interest only or a problem with far-reaching implications?. Vaccine, 19(28-29), pp. 3799–3815.

Fryer, H.R. and McLean, A.R., 2011. Modelling the spread of HIV immune escape mutants in a vaccinated population. PLoS computational biology, 7(12), p. e1002289.

Gandon, S. and Day, T., 2007. The evolutionary epidemiology of vaccination. Journal of the Royal Society Interface, 4(16), pp. 803–817.

Gandon, S., Mackinnon, M. J., Nee, S. & Read, A. F. 2001. Imperfect vaccines and the evolution of pathogen virulence. Nature 414, 751–756.

Gandon, S., Mackinnon, M., Nee, S. & Read, A.F. 2003. Imperfect vaccination: some epidemiological and evolutionary consequences. Proceedings of the Royal Society of London B 270: 1129–1136.

Gog, J.R. and Grenfell, B.T., 2002. Dynamics and selection of many-strain pathogens. Proceedings of the National Academy of Sciences, 99(26), pp. 17209–17214.

Grenfell, B.T., Pybus, O.G., Gog, J.R., Wood, J.L., Daly, J.M., Mumford, J.A. and Holmes, E.C., 2004. Unifying the epidemiological and evolutionary dynamics of pathogens. science, 303(5656), pp. 327–332.

Kennedy, D.A. and Read, A.F., 2017. Why does drug resistance readily evolve but vaccine resistance does not?. Proceedings of the Royal Society B: Biological Sciences, 284(1851), p. 20162562.

Kennedy, D.A. and Read, A.F., 2018. Why the evolution of vaccine resistance is less of a concern than the evolution of drug resistance. Proceedings of the National Academy of Sciences, 115(51), pp. 12878–12886.

Lenski, R.E. and May, R.M., 1994. The evolution of virulence in parasites and pathogens: reconciliation between two competing hypotheses. Journal of theoretical biology, 169(3), pp. 253–265.

May, R.M. and Anderson, R.M., 1983. Epidemiology and genetics in the coevolution of parasites and hosts. Proceedings of the Royal society of London. Series B. Biological sciences, 219(1216), pp. 281–313.

McCarthy, K.R., Rennick, L.J., Nambulli, S., Robinson-McCarthy, L.R., Bain, W.G., Haidar, G. and Duprex, W.P., 2021. Recurrent deletions in the SARS-CoV-2 spike glycoprotein drive antibody escape. Science, 371(6534), pp. 1139–1142.

McLean AR. 1995 Vaccination, evolution and changes in the efficacy of vaccines: a theoretical framework. Proc. R. Soc. Lond. B 261, 389–393.

Mishra, R.P., Oviedo-Orta, E., Prachi, P., Rappuoli, R. and Bagnoli, F., 2012. Vaccines and antibiotic resistance. Current opinion in microbiology, 15(5), pp. 596–602.

Nelson, M.I. and Holmes, E.C., 2007. The evolution of epidemic influenza. Nature reviews genetics, 8(3), pp. 196–205.

Reid, M.C., Peebles, K., Stansfield, S.E., Goodreau, S.M., Abernethy, N., Gottlieb, G.S., Mittler, J.E. and Herbeck, J.T., 2019. Models to predict the public health impact of vaccine resistance: A systematic review. Vaccine, 37(35), pp. 4886–4895.

Roberts, M.G. and Heesterbeek, J.A.P., 2003. Mathematical models in epidemiology (Vol. 215). EOLSS.

Roche, B., Drake, J.M. and Rohani, P., 2011. An Agent-Based Model to study the epidemiological and evolutionary dynamics of Influenza viruses. BMC bioinformatics, 12(1), pp. 1–10.

Rochman, N.D., Wolf, Y.I., Faure, G., Mutz, P., Zhang, F. and Koonin, E.V., 2021. Ongoing global and regional adaptive evolution of sars-cov-2. Proceedings of the National Academy of Sciences, 118(29).

Scherer, A. and McLean, A., 2002. Mathematical models of vaccination. British Medical Bulletin, 62(1), pp. 187–199.

Taubenberger, J.K. and Kash, J.C., 2010. Influenza virus evolution, host adaptation, and pandemic formation. Cell host & microbe, 7(6), pp. 440–451.

van Baalen, M. and Sabelis, M.W., 1995. The dynamics of multiple infection and the evolution of virulence. The American Naturalist, 146(6), pp. 881–910.

Wang, P., Nair, M.S., Liu, L., Iketani, S., Luo, Y., Guo, Y., Wang, M., Yu, J., Zhang, B., Kwong, P.D. and Graham, B.S., 2021. Antibody resistance of SARS-CoV-2 variants B. 1.351 and B. 1.1. 7. Nature, 593(7857), pp. 130–135.

Worby, C.J., Wallinga, J., Lipsitch, M. and Goldstein, E., 2017. Population effect of influenza vaccination under co-circulation of non-vaccine variants and the case for a bivalent A/H3N2 vaccine component. Epidemics, 19, pp. 74–82.

