## Supplementary material for "Evolutionary dynamics of a virus in a vaccinated population": Detailed information about the model used in the main article.

### Modeling an epidemic in an imaginary small town

Burminster is an imaginary small town whose inhabitants experience an outbreak of infectious disease. The people of the town live in houses or apartments, either singly or as couples who may have children at home. The town has several communal institutions: a hospital, a school that admits children of all ages, a nursery for toddlers, a care home for elderly people and a police station. If they do not work in any of these, people work in factories or shops. 'Factories' may be manufacturing companies, warehouses, offices or farms; what they have in common is that they employ workers but do not admit visitors; shops, of course, have both workers and visitors. People travel to work together by public transport. After work they may take their children to a playground or spend the evening with other adults at the cinema or theatre or in a pub.

Once the disease has arrived in the community, anyone may come into contact with infected people and themselves become infected. They may be infected directly, by contact with an infected person, or they may be infected indirectly by contact with surfaces in the home, the workplace or the transit system that have been contaminated by infected people.

As the disease continues to spread, the town authorities may decide to take action. They may decide to close the nursery and school, the non-essential workplaces and shops, or the playgrounds and pubs. They may close them all. They may also advise or enforce restrictions on behaviour, such as enforcing social distancing, the wearing of masks and gloves, or frequent handwashing. If these measures turn out to be effective, the authorities may decide to relax them after a while, but they may be resumed if the disease then begins to flare up again.

The epidemic eventually runs its course and dies out once most people have been infected and either succumbed or recovered and become immune. A complete record of all the infections has been made during the epidemic. This includes how, where and when each case occurred, as well as the history and genealogy of the pathogen itself.

If mutation is allowed, the virus can evolve during the epidemic, either to increase virulence and transmissibility, or to evade the immune response triggered by vaccination or recovery from infection, or both. A complete record of the vaccination status of everyone and the history of the virus population is kept.

#### Detailed account

The program tracks every person in Burminster during their daily activities, perceives when they are at risk of infection and decides whether or not they become infected. Infection and transmission are stochastic events whose course is decided by rules rather than equations. Before anything can happen, you must supply the program with two kinds of information. First, you must supply the values of parameters used in the program, such as the number of houses in the town, or the probability of infection during an encounter with an infected person. Secondly, you must supply demographic information that governs the initial state of the population, such as its age structure or the number of people in different professions.

The program is written in VB6 and you must download the VB6 compiler in order to view, modify and run the code. Some sites from which the compiler can be downloaded are:

http://oceanofexe.com/visual-basic-6-0-download-free/

https://www.raymond.cc/blog/install-visual-basic-6-vb6-in-windows-7-without-microsoft-virtual-machine-for-java/

https://microsoft-visual-basic.en.softonic.com/download

https://onesoftwares.net/visual-basic-6-0/

I have not attempted to use any of these sites. Alternatively, I have supplied an exe version which enables you to run the program, although you cannot see or modify the code.

To operate the exe version of Epidemic Evolution

The exe version is in the zip file which you have downloaded. You should check this executable program for computer viruses. So far as I know, it contains none, but if you decide to run the program I can take no responsibility for any damage to your computer.

First create a directory “c: \windows\desktop” to receive the input and output files.

Create a folder "Epidemic_input_and_output_files" in this directory.

Add two input files to the folder: these specify the age distribution, occupations, case fatality rate and number of children for members of the population. They can be found in the zip file.

Epidemic_DemoDataIn. This specifies the age frequency, marriage frequency, marriage rate, birth rate, natural death rate and case fatality rate for members of the population, at 5-year intervals of age. There are 19 intervals altogether.

Epidemic_ChildrenPerHouseholdIn. This specifies the probability that each married couple have 0, 1, 2 or 3 children living at home, given age of wife.

The output files written by the program are

| | |
|---|---|
| Epidemic_VariablesOut | Values of input parameters |
| Epidemic_PopulateOut | Characteristics of the inhabitants at each address at beginning of run |
| Epidemic_PopulateFinalOut | Characteristics of the inhabitants at each address at end of run |
| Epidemic_WorkPlacesOut | Inhabitants at each place of work |
| Epidemic_WorkRoutesOut | Inhabitants using each route to work |
| Epidemic_CensusOut | Characteristics of population in each cycle, with summary tables |
| Epidemic_DeathRegister | List of inhabitants who die during run |
| VaccineOut | Vaccination history of all members of the population |
| VirusEvolution | History and characteristics of virus strains generated during run |

All these files are in csv format. Note that complete catalogue of virus strains and vaccination status of all individuals will be printed only if requested in final window.

Setting the scene

The parameter values that determine the geography of Burminster and the properties of its inhabitants are supplied through the Object module of the main input Form (ParameterForm). This form provides a console with text boxes in which you write the values that you wish the program to use. There is a default set of parameters already written in these boxes which seem reasonable and give rise to an epidemic that lasts about 200 days, but you can modify them in any way you wish. Here is a list of the parameters that can be set.

One set of parameters defines the human geography of Burminster.

| | |
|---|---|
| TownHouses | The number of townhouses, with separate front doors onto the street. |
| ApartmentBlocks | The number of apartment blocks, with communal entrance and corridors. |
| ApartmentsPerBlock | The (fixed) number of apartments in each block. |
| VacancyRate | The fraction of houses and apartments that are initially vacant. |
| Offices | The number of 'offices', which may be factories, farms, warehouses etc. |
| EssentialOffices | The number (and identity) of offices deemed to be essential. |
| Shops | The number of shops. |
| EssentialShops | The number of shops deemed to be essential. |
| Playgrounds | The number of playgrounds. Only children accompanied by their parents may visit playgrounds. |
| Pubs | The number of places of public resort: theatres, cinemas, bars, pubs etc. Only adults may visit these. |
| Routes | The number of routes by which people may travel to their workplace or to the nursery, school, hospital or care home. |
| HospitalBeds | The number of beds available in the hospital. |
| CareHomeRooms | The number of rooms in the care home. |

Another set governs the process of infection and its time course.

An option button determines whether both direct and indirect infection are allowed, or direct infection only. If the virus is allowed to evolve, only direct infection should be allowed.

| | |
|---|---|
| DirectInfection | The probability per event of becoming infected through an encounter with an infected person. |
| IntimateDirectInfection | The corresponding but greater probability of becoming infected through intimate contact between doctors and patients in the hospital or between residents and staff in the care home. |
| LimitDirectInfection | The limiting (maximal) probability of infection from a family member at home |
| IndirectInfection | The probability per event of becoming infected by touching a contaminated object. |
| Nobjects | The number of objects at a workplace or on a route that may be contaminated when touched by an infected person. |
| Ntouch | The number of these objects that is touched by any given person on any given day. |
| Encounters | The number of other people encountered each day in the workplace or when travelling to it. |
| IncubationPeriod | The number of days between becoming infected and the onset of symptoms. |
| SymptomaticPeriod | The number of days during which symptoms of the disease are expressed, without becoming severe enough to cause death. |
| CrisisPeriod | The number of days during which symptoms are sufficiently severe to cause death. Any person who survives the crisis period recovers and is immune. |

| | |
|---|---|
| Persistence | The number of days for which the pathogen survives on contaminated surfaces. |
| TransitionSevere | The probability per day of transition from symptomatic to severe disease. |
| TransitionDeath | The probability per day of transition from severe disease to death, for people who have been hospitalized. |
| TransitionDeathAtHome | The corresponding but greater probability for sick people who remain at home, usually because there are no beds available at the hospital. |

There are also parameters which describe the general health of each person.

| | |
|---|---|
| FallSick | The probability per day of falling sick from causes other than the disease. |
| GetWorse | The probability per day that a sick person will become critically ill. |
| Die | The probability per day that a critically ill person will die. |
| CriticalPeriod | The length of time in days that a person is critically ill; if they survive as long they recover and remain alive but in poor health. |

You then supply some demographic data to set up the initial population of Burminster. This is held in two files, both written in csv format. These files are held in a folder and their location and names are specified by the user in the text box of the InputOutputForm. You should construct this folder with the input and output files before running the program. The output files are named at the end of this document, but you can make up your own if you wish. Once the files are correctly specified in the FileInputBox of the InputOutputForm, click twice to proceed to the next form.

The first input file (Epidemic_DemoDataIn) gives basic demographic data in five-year categories of age.

| | |
|---|---|
| AgeFreq | The frequency of each five-year age group in the population. |
| MarriageFreq | The fraction of people in this age group who are married (i.e. man and woman living as a couple in the same house). |
| MarriageRate | The probability that a person of given age will marry. (This is only called in multi-year runs.) |
| BirthRare | The probability that a woman of given age will give birth in the current year. |
| DeathRate | The probability that a person of given age will die in the current year. |
| CFR | The case fatality rate for the disease: the probability that a person of given age will die from contracting the disease. |

The second file (Epidemic_ChildrenPerHouseholdIn) gives the number of children per household as a function of the age of the female resident of a house.

The rates of marriage, birth and death are called only in multi-year runs and are not used in this version. No distinction of gender is made for age structure (although one exists). No more than three children per household are permitted (for convenience). The default values are based on data for Britain and Canada, but they are not intended to represent any particular country and merely ensure that the demography is not wholly unrealistic. The values can be changed at will by altering the input files.

#### The initial population

The demographic data are now used to populate the houses and apartments of the town. Each is vacant or holds one or two adults. Two adults living together are assigned children on the basis of their age, according to the schedules in the two input files. Thus, any house or apartment may have up to five occupants. Each occupant is assigned a permanent name (actually a number) and a list of features.

| | |
|---|---|
| Gender | Male or Female with equal probability. |
| Age | Age in years, with probability from input demographic data. |
| Job | Occupation, with probability set by recent UK government data.<br>ChildMinder (works in Nursery)<br>Teacher (includes all educational staff; works in School)<br>Doctor (includes all medical staff; works in Hospital)<br>ElderCarer (works in Care Home)<br>Police (works in Police Station)<br>Office worker (works in factory, warehouse, office, farm etc)<br>Shop assistant (works in Shop)<br>Homeworker (not in paid employment; works at home).<br>Retired (all persons over 65 years of age are assumed to have retired). |
| Health | Health status: Robust or Fragile. |
| Infected | Whether or not infected (all persons are initially uninfected). |
| Immune | Whether or not immune (having been vaccinated or recovered from disease). |
| Hospitalized | Whether or not hospitalized (because disease symptoms are severe or fragile health has deteriorated to critical illness). |
| Marital status | Single or married; if married, the name of their spouse is attached. |

The Care Home is then populated by moving anyone over 80 years of age (or a couple of which one or both are over 80) into a room in the Care Home, if one is available. All hospital beds are initially empty.

The arrival of the disease

The epidemic begins when a single person, chosen at random, becomes infected. It may die down almost immediately, especially if the disease is not very infectious. Otherwise it will spread through the population of the town. Immigration from other towns is not permitted in this version.

The daily round

The day begins at home. All adults in paid employment then travel by a particular route to their workplace, where they spend the day. Toddlers travel to the Nursery and children to the School. Adults travel to an Office or Shop, or work at home, or are retired and do not work. After work, people may visit the playground or the pub. Whether at home, travelling to work, working or playing, they will encounter a specified list of other people: the people living in the same house, travelling on the same route, working in the same place, visiting the same shop or resorting to the same playground or pub. These daily activities pose a risk of infection, either directly from one person to another, or indirectly by touching contaminated surfaces, if this is permitted.

**Direct transmission**. Each activity involves meeting other people. Anyone engaged in a given activity meets a certain number of other people and may be directly infected by contact with them. This list of contacts includes fellow travellers on a given route, customers in a given shop, people in good health visiting hospitalized friends, people visiting their elderly parents in the care home, and so forth.

**Indirect transmission**. Each activity also involves contact with contaminated surfaces. You can specify by an option button whether or not indirect infection takes place. The level of contamination depends on the number of infected people who participate in that activity. It is evaluated as follows.

The number of infected people using a route or workplace is Tinfected.

The number of objects on a route or at a workplace that might be infected is Nobjects. These might be poles, handles, seat covers etc.

The number of objects that a person touches is Ntouch.

An object touched by an infected person becomes contaminated. It is fully contaminated, with the same level of contamination no matter how many infected people touch it.

Contamination persists for a certain number of days and then disappears completely.

If there is a single infected person who touches Ntouch random objects then

Prob(object contaminated) = Ntouch/Nobjects = Pcontaminated

Prob(not contaminated) = 1 – Pcontaminated.

With Tinfected infected people, the probability that an object is not contaminated is (1-Pcontaminated)^Tinfected.

Hence the probability that the object is contaminated is 1 – (1-Pcontaminated)^Tinfected.

The number of contaminated objects touched is then

Ncontaminated = Ntouch[1 – (1-Pcontaminated)^Tinfected]

The probability of infection from touching a contaminated object is (the input parameter) Q. The probability of not being infected is then 1 – Q.

The probability of not being infected by touching Ncontaminated contaminated objects is (1- Q)^Ncontaminated.

Hence the probability of being infected by touching Ncontaminated contaminated objects is 1 – (1-Q)^Ncontaminated.

Hence the probability of infection per day during a journey or at the workplace is

ProbInfection = (1-Q)^[[1-(1-Pcontaminated)^Tinfected}]

This is the function used to assess the probability of indirect infection.

At the end of the day's activities, everyone's health status is examined. Some people may have become critically ill, from causes unrelated to the epidemic, and will be moved to the hospital if beds are available. Others will be progressing through the stages of infection, and may present severe symptoms which require hospitalization, again if beds are available. Some may die, and their names are removed from the house or apartment where they used to live, and deleted from the lists of activities they once participated in.

Anyone who survives the disease becomes immune, either permanently or temporarily. If temporarily, the period of immunity should be specified.

#### Intervention

As the epidemic progresses, the town authorities may decide to intervene in an attempt to slow the spread of the disease. The next stage in the program is another console, which looks rather like the parameter input console except that some of the boxes are concealed whereas new ones have appeared. There is an option button so that you can choose to "Do nothing". Otherwise, the measures that the authorities may take are specified by ticking the new check boxes.

First, close some or all public and private institutions.

Close the nursery and school;
Close the hospital and care home to visitors;
Close playgrounds and pubs;
Close public transport;
Close non-essential factories and shops.

Secondly, enforce some patterns of social behaviour.

Social distancing and banning of large groups (reduces number of encounters with other people);
Wear masks (reduces probability of direct infection per encounter);
Wear gloves (reduces number of contaminated objects touched);
Wash hands (reduce probability of infection from touching contaminated object);
Cleaning public spaces and transport (reduces time that pathogen persists on surfaces).

Thirdly, people showing symptoms of disease can be placed in quarantine. This quarantine may be extended to family members or to the people they work with.

Some people might not comply with these regulations, and you can designate the percent of people who will comply with behavioural restrictions or quarantine. These are presumably enforced by the police, but I haven't provided for that yet.

Whatever pattern of intervention is decided on, it must be imposed at some point, depending on the progress of the epidemic. There are three choices. (These are presented as check boxes but should be treated as option buttons; choose only one.)

Early: when the there is a consistent increase in the number of cases for the specified period.
Late: the same, for twice the specified period.
Too late: the same, for four times the specified period.

It would be unrealistic to suppose that the authorities can react instantly to changes in disease incidence, and you can specify the time lag in the "Decision latency" box. For example, if you specify 7 (days) for Early intervention then the number of new cases must increase every day for seven consecutive days before the authorities will intervene.

Once an intervention has been put in place, it must be lifted sooner or later, and this decision, like the decision to impose it in the first place, will depend on the course of the epidemic. There are again three choices.

> Early: when the number of new cases is less than the peak number (for the specified number of days).
> Late: the same, for twice the specified number of days.
> Never: keep up all the measures until almost everyone is immune.

Early and Late relaxation are subject to the same decision latency as imposing the restrictions in the first case. If you would like to be able to re-impose restrictions if the disease flares up again, tick the check box labelled “Resume if necessary”; the same criteria for beginning and ending restrictions will be applied.

The measures that have been chosen may be applied and withdrawn more or less gradually. You have three choices.

> Abrupt: all restrictions are imposed or relaxed abruptly (on the same day).
> Gradual: Nursery, school, factories and shops re-open immediately, but playgrounds and pubs remain closed and behavioural regulations and quarantine in force for another month.
> Slow: nursery and school open immediately; factories and shops open after one month; playgrounds and pubs open, and behavioural regulations and quarantine cease, only after another three months.

<u>Virus genealogy</u>

If only direct person-to-person transmission is allowed, the complete chain of infection can be traced. This enables us to calculate the average number of new infections that each infected person gives rise to. This is the basic reproduction number $R_0$ of the virus, and a basic theoretical result is that the epidemic will spread if $R_0 > 1$. In empirical examples the reasoning may be circular, with $R_0$ estimated from the rate of spread of the virus. In the Epidemic program $R_0$ is calculated directly from the observed chains of infection. The disease may spread even if $R_0 < 1$ when indirect infection is allowed.

Suppose that the rank of a host is k if it is the k-th susceptible person to be infected by contact with a given infected person. The descent of the particular virus population that infects a given person can then be expressed in terms of the ranks of its ancestral populations: thus, 1.2.2 would indicate that this population represents the second successful transmission (2) from a population that itself was also the second successful transmission (2.2) from the first successful transmission of the founding population (1.2.2). This notation denotes a unique path of descent for each virus population and enables the entire tree of descent to be recovered.

Alternatively, the list of ancestors by permanent name will be generated for each infection. Virus pedigrees are printed to file if the box “Print complete virus ancestry’ on MainForm is ticked.

This procedure fails if indirect transmission is allowed, because the ancestry of a new infection cannot be ascertained (unless, in principle, the route or place is known to have been contaminated by a single identified infected person). If an infection has been acquired indirectly its ancestry is denoted “0” and instead of a single continuous tree the pattern of descent becomes a series of short isolated chains.

Virus genetics

**Loci.** Two genetic loci govern the interaction between virus and host. Each consists of a string of bits (0.1) of specified length. The Phenotype locus (P-locus) governs the proliferation of the virus within the host, and thereby the virulence and transmissibility of a given strain. The ancestral strain has specified values of virulence and transmissibility for naïve (never infected), recovered (from infection) and vaccinated hosts. The Immunity locus (M-locus) governs the immune response through the complementarity of the sequence at this locus with sequences (if any) held in the immune memory of a host. Naïve unvaccinated hosts have no immune memory and are thus universally susceptible to all strains of virus.

**Mutation.** If mutation is permitted at either locus it occurs with specified probability per locus. Should it occur, the modified bit is chosen at random and shifted 0 → 1 or 1 → 0. Mutation at the M-locus directly modifies the immune properties of the virus. Mutation at the P-locus always increases virus titre, and thereby increases both virulence and transmissibility. The degree of modification is a random increment between the original value before mutation and a specified upper limit (implying that on average it increases asymptotically from the ancestral value towards a fixed upper limit). The increment is independently drawn for virulence and transmissibility (which will nevertheless be positively correlated, since both increase). The fixed upper limit is specified separately for naïve, recovered and vaccinated individuals.

**Recombination.** If coinfection is allowed, any given infected individual may receive, with specified probability, a second strain of the virus. A random individual is chosen to supply the coinfecting strain. If the two strains have previously acquired a mutation at either the P-locus or the M-locus or both then the P-locus of the coinfecting strain is substituted for the P-locus of the resident strain. Recombination is always intergenic; intragenic recombination is not permitted.

<u>Host immune memory.</u>

In response to infection by a strain of virus with a given epitope sequence at its M-locus, the host generates an antibody with the complementary sequence and stores this in its immune memory. For example, if the virus M-locus is 001011001, the host generates and stores 110100110. If the host survives and is subsequently exposed to the same strain of virus, it is able to express the stored sequence and may thereby disable the virus. If it is subsequently exposed to a different strain, its immune memory and the M-locus of the virus will be imperfectly complementary. For example, if the host has stored 110100110 whereas the virus M-locus sequence is 101011001 then the degree of complementarity is only 9 rather than the full 10. This may or may not be sufficient to trigger an immune response: the effectiveness of the immune response depends on how the clearance of the virus is related to the degree of complementarity. When a host individual is exposed to infection, the M-locus of the virus is compared with each sequence in the host immune memory, and the sequence in memory with the greatest complementarity is chosen. The function that has been specified to relate complementarity to immunity is then used to calculate the probability that the host individual is immune to the virus strain. Three functions are supported:

> Threshold. The host is immune if the sequence in memory with greatest complementarity equals or exceeds a specified threshold. The parameters are the threshold value, and the probability of being immune if this threshold is met or exceeded.
>
> Accelerating. The probability of immunity decreases exponentially as complementarity decreases. The parameters are the probability of immunity when complementarity is complete, and the slope of the exponential decline relating the probability of immunity to complementarity.
>
> Decelerating. The inverse relation, when the probability of immunity increases asymptotically with complementarity.

The parameters for each model of immunity may have different values for naïve, recovered and vaccinated hosts. These parameters define the breadth and efficacy of the vaccine.

<u>Vaccination</u>

A vaccinated individual holds the sequence complementary to the ancestral virus strain in its immune memory. The vaccination program begins on a specified date, and is delivered to three cohorts at specified intervals. The cohorts are defined by age and occupation. The standard parameter set specifies: Cohort 1, all people greater than 75 years of age, and care-home workers; Cohort 2, all people greater than 65 years of age, and health-care workers and teachers; Cohort 3, everyone else. Any other schedule can be specified if desired. Vaccination begins 6 weeks after the first case and proceeds in cohorts separated by three weeks. People are vaccinated regardless of infection status. Vaccination may be compromised in several ways. First, compliance may be incomplete, so that only a fraction of the population is vaccinated. Secondly, the vaccination process itself may fail, so that someone who has been vaccinated in fact receives no protection. Thirdly, the efficacy of the vaccine may lapse after a given length of time.

Output

You can follow the course of the epidemic by opening the Immediate window, which gives a day-to-day summary and some information about the final outcome. There are currently eight output files which are printed to the folder you have designated.

Parameter values. The values used to run the program are recorded in the file "Epidemic_VariablesOut.csv".

Initial population. The characteristics of the initial population are recorded in the file "Epidemic_PopulateOut.csv".

Workplaces. The identity of people working in different workplaces is recorded in the file "Epidemic_WorkPlacesOut.csv".

Routes. The identity of people travelling by different routes is recorded in the file "Epidemic_WorkRoutesOut.csv".

Course of epidemic. The progress of the epidemic is recorded each day in the file "Epidemic_CensusOut.csv", which also contains a listing of where each case occurred and a summary of virus reproduction. A complete listing of the descent of each infection will be printed in the box on EpidemicMainForm is checked.

Deaths. The identity of all people dying during the epidemic, whether from disease or from other causes, is recorded in 'DeathRegister'.

Virus evolution. The frequency of strains is recorded at the end of each day in 'VirusEvolution', with a summary of the characteristics of each strain written at the end of the run. Note that new strains may bear the same identification number as extinct strains (in order to save memory).

Vaccination. The vaccination history of every individual in the population is recorded in 'VaccineOut'.

Others can be added by modifying the code.

**Default parameters**. The program as supplied allocates parameter values (in ParameterForm) that usually (but not always, depending on the random number seed) produce an epidemic in a population of about 4,000 people. These can be altered at will. Increasing the population size may cause an 'Out of Memory' error, since the dimensions specified for array variables are rather generous.
